# Evaluation of anti-spike glycoprotein antibody and neutralizing antibody response of different vaccine platforms. A protocol of systematic review and meta-analysis of COVID-19 vaccine clinical trial studies

**DOI:** 10.1101/2022.05.18.22275247

**Authors:** Mohammad Mirzakhani, Safa Tahmasebi, Mohammadreza Dashti, Mohammad Reza Mohammadi, Manizhe Faghih, Mousa Mohammadnia-Afrouzi, Hadi Esmaeili Gouvarchin Ghaleh, Jafar Amani

## Abstract

**Background:** The severe acute respiratory syndrome coronavirus 2 (SARS-CoV-2) emerged in late 2019 and spread globally, prompting an international effort to accelerate development of a vaccine. SARS-CoV-2 transmit among the people fast and infected thousands of people daily around the world. Because of rapid transmission of SARS-CoV-2 among the people, there is an urgent need to prevent people from infection or hospitalization and control the disease.

**Methods:** We will search electronic databases such as PubMed, Web of Science, Cochrane (CENTRAL), Scopus, Google scholar, the key journals (vaccine and vaccines). Moreover, trial registry including clinicalTrials.gov, WHO ICTRP, and ISRCTN will be searched. We will only select all clinical trial studies in any phases of evaluation (i.e. phase I, II, II, IV). For anti-spike glycoprotein antibody (IgG) response and neutralizing antibody response, we will report Ratio of Geometric Mean (RoGM), Ratio of Mean (RoM) or standardized mean difference (SMD) depends on type of articles.

**Discussion:** Various vaccine platforms have been developed to increase the resistance to the SARS-CoV2 virus and reduce hospitalization and mortality rates. The comprehensive data gathering and analysis of results will guide scientists about the best available evidence. Moreover, the current study results may indicate which of the vaccine platforms are more effective and safe for COVID-19.

## Background

The outbreak of new coronavirus disease-2019 (COVID-19) due to RNA virus severe acute respiratory syndrome coronavirus 2 (SARS-CoV-2) affected millions of people worldwide. SARS-CoV-2 is one of the Beta coronavirus strains which was identified in Wuhan, China in December 2019 for the first time (1-3). After entering the body, SARS-CoV-2 binds to its receptor via the 180-KD spike protein (4-6). The studies showed that the angiotensin-converting enzyme-2 (ACE-2) on host cells acts as a high-affinity receptor for SARS-CoV-2 (7-10). Based on the world health organization statistics, until march 2022, the number of individuals infected with SARS-CoV-2 was over than 450 million people and the number of death due to COVID-19 reached over than 6 million. Patients showed the different severity of disease including asymptotic, mild, moderate, severe, and critical disease (11-13). SARS-CoV-2 transmit among the people fast and infected thousands of people daily around the world (14). Because of rapid transmission of SARS-CoV-2 among the people, there is an urgent need to prevent people from infection or hospitalization and control the disease. For this purpose, candidate vaccines against SARS-CoV-2 were developed and several of them evaluated the pre-clinical and clinical studies with published articles(15-18). There are several vaccine platforms against SARS-CoV-2 with different adverse reactions, immunogenicity, and efficacy(19-24). In this systematic review and meta-analysis study, we aim to evaluate and compare the anti-spike glycoprotein antibody and neutralizing antibody response of different vaccine platforms as well as their adverse reactions. To our knowledge there is no systematic review and meta-analysis to evaluate and compare the safety and efficacy of different vaccine platforms with the mentioned primary and secondary outcomes as well as subgroup analyses.

## Objective

### Primary objectives

1. Evaluation and comparison of anti-spike glycoprotein antibody (IgG) response of different vaccine platforms after two doses of vaccine
2. Evaluation and comparison of neutralizing antibody response of different vaccine platforms after two doses of vaccine

These outcomes will be analyzed separately based on different vaccine platforms and different age groups 18-59 years and ≥ 60 years if the studies reach to adequate number (≥4).

### Secondary objectives

1. Occurrence of any adverse reactions within 7 days and overall adverse reactions within 28 days after vaccination Studies with data of adverse reactions within 14 days will analyze with those with adverse reactions within 7 days together.
2. Evaluation and comparison of the IFN-γ-producing TH1 cells in different vaccine platforms provided that inclusion of enough studies
3. Incidence of seroconversion after the first and the second doses of vaccine separately
4. Incidence of symptomatic/asymptomatic cases and severe/hospitalized cases

These outcomes will be analyzed separately based on different vaccine platforms and different age groups 18-59 years and ≥ 60 years if the studies reach to adequate number (≥4).

### Subgroup analyses

1- Performing subgroup analysis based on different vaccine platforms In the initiation of analysis, we will not have all study combination and will perform subgroup analysis based on different vaccine platforms.
5- Performing subgroup analysis based on age groups 18-59 years and ≥ 60 years In the different vaccine platforms subgroups, we will have subgroup analysis based on age groups 18-59 years and ≥ 60 years.
6- Performing subgroup analysis based on gender variable This subgroup analysis will be performed in the each age groups 18-59 years and ≥ 60 years separately.
7- IgG against any parts of spike glycoprotein will analyze together and if studies are adequate, we will perform subgroup analysis based on IgG against different parts of spike glycoprotein i.e. RBD and etc.
8- Studies with nature of single dose and with the nature of two doses will analyze together
9- Performing subgroup analysis after single dose and after two doses of vaccine for primary outcomes, in the studies with the nature of two doses of vaccine, if enough studies are available.

## Method

### Eligibility criteria

#### Type of study

We will only select all clinical trial studies in any phases of evaluation (i.e. phase 1, 2, 3, 4). The studies will include if they have the data of vaccination group and placebo/control group, if they have one of our primary or secondary outcomes data, and if they have heterologous vaccination design with the same vaccine platforms. The studies will not include if they have not control group or have a convalescent group as a control or as a comparison group, if they have not any of our primary or secondary outcomes, and if they have not heterologous vaccination design with the same vaccine platforms. The cohort, and animal studies will be excluded. The pre-print repositories will not be searched but pre-print studies which are extracted from mentioned databases will include in this study.

#### Type of participant

We will enroll the following participants in this systematic review and meta-analysis. The healthy participants that received one dose or two doses of vaccine will include if they have one of our primary or secondary outcomes data. The healthy participants that received one dose or two doses of placebo will include if they have one of our primary or secondary outcomes data. The immunocompromised patients, transplant recipients, patients with cancer, and so on will not include in this study. The participants will be any age, any gender, and any demographic data.

### Type of outcome measurements

#### Primary outcomes measurements

For anti-spike glycoprotein antibody (IgG) response and neutralizing antibody response, we will report Ratio of Geometric Mean (RoGM) and/or Ratio of Mean (RoM).

#### Secondary outcomes measurements

1. For the occurrence of any adverse reactions within 7 days and overall adverse reactions within 28 days after vaccination, we will report odds ratio (OR)/risk ratio (RR). Moreover, the prevalence of mentioned adverse reactions will be reported.
2. For the frequency of IFN-γ-producing TH1 cells in different vaccine platforms, we will report standardized mean difference (SMD) or RoM.
3. For the incidence of seroconversion after the first and the second doses of vaccine, we will report the prevalence of seroconversion in the vaccinated and placebo participant separately.
4. For the incidence of symptomatic/asymptomatic cases and severe/hospitalized cases, we will report prevalence of symptomatic/asymptomatic cases and severe/hospitalized cases in the vaccinated and placebo groups, separately.

### Search strategy

#### Source of primary researches

We will search several sources without language limitation to achieve the goal of research. The sources include electronic database such as PubMed, Web of science, Cochrane (CENTRAL), Scopus, Google scholar, the key journals (vaccine and vaccines). Trial registry including clinicalTrials.gov, WHO ICTRP, and ISRCTN will be searched. In addition, “COVID-19 vaccine tracker and landscape” of WHO, “Cochrane vaccine mapping tool”, and the reference list of included primary will be searched.

#### Search time interval

Primary studies will search from 1 January 2020 up to 30 September 2021.

### Key words and syntax

The main search terms are “SARS-CoV-2 vaccine”, vaccine, “COVID-19 vaccine” and COVID-19 and “Coronavirus disease 2019” that will be searched in MeSH databases. We also will use free text method to achieve a great syntax. The generic syntax in PubMed database is in the below.

(“SARS-CoV-2 vaccine” OR vaccine OR “ChAdOx1 nCoV-19” OR AZD1222 OR ChAdOx1 OR ChAdOx1-S OR “recombinant adenovirus type-5-vectored” OR “recombinant adenovirus type” OR recombinant OR “COVID-19 vaccine” OR “non-replicating adenovirus” OR Ad5-vectored OR CanSino OR “Ad5 vectored” OR “vector-based COVID-19 vaccine” OR rAd26 OR rAd5 OR “Recombinant viral vector-based vaccines” OR “viral vector” OR Ad26.COV2.S OR Ad26 OR Ad5 OR vector OR NVX-CoV2373 OR “rSARS-CoV-2 vaccine” OR “recombinant nanoparticle vaccine” OR “nanoparticle vaccine” OR “Inactivated SARS-CoV-2 Vaccines” OR “Inactivated SARS-CoV-2 Vaccine” OR inactivated OR “Inactivated COVID-19 Vaccines” OR “Inactivated Vaccines” OR “Inactivated Vaccine” OR “whole-virus COVID-19 vaccine” OR “COVID-19 candidate vaccines” OR BBIBP-CorV OR “inactivated virus” OR CoronaVac OR “mRNA-1273 SARS-CoV-2 Vaccine” OR mRNA-1273 OR “mRNA-1273 vaccine” OR “mRNA vaccine” OR “candidate SARS-CoV-2 vaccines” OR “Covid-19 Vaccine Candidates” OR “Covid-19 Vaccine” OR replicating OR non-replicating OR BNT162b1 OR BNT162b2 OR “COVID-19 RNA vaccine” OR “RNA vaccine” OR “RNA-based SARS-CoV-2 vaccine” OR “protein subunit” OR ZF2001 OR “receptor-binding domain” OR EpiVacCorona OR Virus-Like-Particles OR “Virus-like particles” OR “Virus-like particle” OR “subunit vaccine”) AND (“COVID-19” OR “Coronavirus disease 2019” OR “SARS Coronavirus 2” OR “Coronavirus disease 19” OR 2019-nCoV OR “Wuhan coronavirus” OR “2019 novel coronavirus” OR “COVID-19 virus” OR “coronavirus disease 2019 virus” OR “COVID19 virus” OR “COVID-19 pandemic” OR “severe acute respiratory syndrome coronavirus 2” OR SARS-CoV-2 OR “SARS-CoV-2 Virus” OR “acute respiratory failure” OR “acute respiratory distress syndrome” OR ARDS OR “intensive care unit” OR “intensive care” OR mortality OR outbreak OR Severe/critical OR Mild/moderate OR prevalence) AND (randomized controlled trial[pt] OR controlled clinical trial[pt] OR randomized[tiab] OR placebo[tiab] OR drug therapy[sh] OR randomly[tiab] OR trial[tiab] OR groups[tiab] NOT animals[mh] NOT humans[mh]) AND 2020/01/01:2022/03/30[dp]

### Study selection

We will export our search output into End-note software and then duplicated primary studies will be deleted (only one version of primary studies will be kept). Screening step (included/probable included versus excluded primary researches) will be performed according to title and abstract. Then two reviewers using full text will independently conduct selection/confirmation process (included versus excluded primary researches) according to eligibility criteria. Any discordance/disagreement will be resolved by consensus otherwise opinion of third subject expert will be considered.

### Risk of bias assessment

Two reviewers independently will conduct quality assessment of primary studies by using checklist of quality assessment for interventional studies (Cochrane risk-of-bias tool). Any disagreement will be resolved by discussion/consensus otherwise opinion of third reviewer will be considered.

### Data extraction and data analysis

#### Data extraction

Data extraction form will be developed by using included papers. Two reviewers independently extract information by means of data extraction form and discordance will be resolved by consensus or opinion of third reviewer. If there were incomplete data, we will contact with corresponding authors of studies.

The following data will be extracted from the primary studies:

Authors’ name, publication year, gender, anti-spike glycoprotein antibody geometric mean titre (GMT)/geometric mean concentration (GMC), neutralizing antibody GMT, median (IQR) for anti-spike glycoprotein antibody or neutralizing antibody, number of cases with adverse reactions within 7 days/28 days in the control and vaccination groups, median (IQR) for IFN-γ-producing TH1 cells, number of cases with seroconversion and total number of cases, number of cases with symptomatic/asymptomatic features/with severe/hospitalized features and dosage of vaccines.

#### Data analysis

First of all, we will select the concordant valid effect size (mentioned in the Type of Outcome Measurements section) and then in according to the methodological similarities between final eligible papers the appropriate combination model––fixed-effect model or random-ffect model––will be selected and combined effect size will be plotted using forest plot.

We will assess statistical heterogeneity of studies using I^2^ measure and if there was any heterogeneity, we will try to find potential source of heterogeneity using subgroup analysis (for more than 3 studies) or meta-regression (for less than 4 studies). The severe heterogeneity will be considered as I^2^ >=50%.

We will use Funnel plots and Begg’s and Egger’s tests to assess publication bias and sensitivity analysis will be performed by one-out remove method.

### Certainty of evidence

We will assess the certainty of the evidence using the Grading of Recommendation Assessment, Development and Evaluation (GRADE) tool.

## Discussion

Various vaccine platforms have been developed to increase the resistance to the SARS-CoV2 virus and reduce hospitalization and mortality rates (25-27). Given the critical need for pandemic control, efforts have been made to formulate high-performance vaccines with the maximum effective immune response and minimal side effects (28, 29). So far, various prophylactic vaccines with different efficacy and safety have been approved for the COVID-19, depending on their structure and ability to elicit immune responses (30-33). Multiple preclinical and clinical trial studies have been conducted or are underway to examine various aspects of COVID-19 vaccine platforms (15, 17, 34). However, there are differing views on the efficacy and safety of these platforms based on the reported results of different studies (35, 36). Therefore, in a current systematic review and meta-analysis study, for the first time, we will comprehensively search all databases, collect and analyze the related data to antibody responses (anti-spike glycoprotein antibody (IgG) and neutralizing antibody responses) and side effects of different anti-SARS-CoV2 vaccine platforms.

The comprehensive data gathering and analysis of results will guide scientists about the best available evidence. Moreover, the current study results may indicate which of the vaccine platforms are more effective and safe for COVID-19. This protocol describes our objectives and methodology for the meta-analysis of the primary interventional studies. We will search and select all clinical trial studies in any phases of evaluation (i.e., phases 1, 2, 3, 4). The studies will include whether they have the data of vaccination group and placebo/control group and have one of our primary or secondary outcomes.

As the strengths of this study, we will search several sources without language limitation to achieve the research goal. The sources include electronic databases such as PubMed, Web of Science, Cochrane Central Register of Controlled Trials (CENTRAL), Scopus, Google Scholar, the key journals (vaccine and vaccines). Trial registries, including ClinicalTrials.gov, WHO ICTRP, and ISRCTN, will be searched. In addition, “COVID-19 vaccine tracker and landscape” of WHO, “Cochrane vaccine mapping tool”, and the reference list of included primary will be searched. Also, the publication bias of primary studies will be evaluated. Eventually, we expect that our study notably advances the knowledge about the safety and efficacy of various COVID-19 vaccine platforms.

## Data Availability

This is a protocol study with no data or results

## References

1. Pal M, Berhanu G, Desalegn C, Kandi V. Severe Acute Respiratory Syndrome Coronavirus-2 (SARS-CoV-2): An Update. Cureus. 2020;12(3):e7423–e.

2. Hu B, Guo H, Zhou P, Shi Z-L. Characteristics of SARS-CoV-2 and COVID-19. Nature Reviews Microbiology. 2021;19(3):141–54.

3. Liu Y-C, Kuo R-L, Shih S-R. COVID-19: The first documented coronavirus pandemic in history. Biomedical Journal. 2020;43(4):328–33.

4. Veenstra TD, Pauley B, Injeti E, Rotello RJ. &lt;em&gt;In vitro&lt;/em&gt; Characterization of SARS-CoV-2 Protein Translated from the Moderna mRNA-1273 Vaccine. medRxiv. 2022:2022.03.01.22271618.

5. Zhang Q, Xiang R, Huo S, Zhou Y, Jiang S, Wang Q, et al. Molecular mechanism of interaction between SARS-CoV-2 and host cells and interventional therapy. Signal Transduction and Targeted Therapy. 2021;6(1):233.

6. Dejnirattisai W, Zhou D, Ginn HM, Duyvesteyn HME, Supasa P, Case JB, et al. The antigenic anatomy of SARS-CoV-2 receptor binding domain. Cell. 2021;184(8):2183–200.e22.

7. Ni W, Yang X, Yang D, Bao J, Li R, Xiao Y, et al. Role of angiotensin-converting enzyme 2 (ACE2) in COVID-19. Critical Care. 2020;24(1):422.

8. Zamorano Cuervo N, Grandvaux N. ACE2: Evidence of role as entry receptor for SARS-CoV-2 and implications in comorbidities. Elife. 2020;9:e61390.

9. Zhang H, Penninger JM, Li Y, Zhong N, Slutsky AS. Angiotensin-converting enzyme 2 (ACE2) as a SARS-CoV-2 receptor: molecular mechanisms and potential therapeutic target. Intensive Care Med. 2020;46(4):586–90.

10. Yang J, Petitjean SJL, Koehler M, Zhang Q, Dumitru AC, Chen W, et al. Molecular interaction and inhibition of SARS-CoV-2 binding to the ACE2 receptor. Nature Communications. 2020;11(1):4541.

11. Gao Z, Xu Y, Sun C, Wang X, Guo Y, Qiu S, et al. A systematic review of asymptomatic infections with COVID-19. J Microbiol Immunol Infect. 2021;54(1):12–6.

12. Li Y, Shi J, Xia J, Duan J, Chen L, Yu X, et al. Asymptomatic and Symptomatic Patients With Non-severe Coronavirus Disease (COVID-19) Have Similar Clinical Features and Virological Courses: A Retrospective Single Center Study. Frontiers in Microbiology. 2020;11.

13. Zhu Z, Cai T, Fan L, Lou K, Hua X, Huang Z, et al. Clinical value of immune-inflammatory parameters to assess the severity of coronavirus disease 2019. International Journal of Infectious Diseases. 2020;95:332–9.

14. Sender R, Bar-On YM, Gleizer S, Bernshtein B, Flamholz A, Phillips R, et al. The total number and mass of SARS-CoV-2 virions. Proceedings of the National Academy of Sciences. 2021;118(25):e2024815118.

15. Kyriakidis NC, López-Cortés A, González EV, Grimaldos AB, Prado EO. SARS-CoV-2 vaccines strategies: a comprehensive review of phase 3 candidates. npj Vaccines. 2021;6(1):28.

16. Krammer F. SARS-CoV-2 vaccines in development. Nature. 2020;586(7830):516–27.

17. Ahmed S, Khan S, Imran I, Al Mughairbi F, Sheikh FS, Hussain J, et al. Vaccine Development against COVID-19: Study from Pre-Clinical Phases to Clinical Trials and Global Use. Vaccines. 2021;9(8):836.

18. Chakraborty S, Mallajosyula V, Tato CM, Tan GS, Wang TT. SARS-CoV-2 vaccines in advanced clinical trials: Where do we stand? Advanced Drug Delivery Reviews. 2021;172:314–38.

19. Khoshnood S, Arshadi M, Akrami S, Koupaei M, Ghahramanpour H, Shariati A, et al. An overview on inactivated and live-attenuated SARS-CoV-2 vaccines. Journal of Clinical Laboratory Analysis.n/a(n/a):e24418.

20. Nagy A, Alhatlani B. An overview of current COVID-19 vaccine platforms. Computational and Structural Biotechnology Journal. 2021;19:2508–17.

21. Rahman MM, Masum MHU, Wajed S, Talukder A. A comprehensive review on COVID-19 vaccines: development, effectiveness, adverse effects, distribution and challenges. Virusdisease. 2022:1–22.

22. He Q, Mao Q, Zhang J, Bian L, Gao F, Wang J, et al. COVID-19 Vaccines: Current Understanding on Immunogenicity, Safety, and Further Considerations. Front Immunol. 2021;12.

23. Peng X-L, Cheng J-S-Y, Gong H-L, Yuan M-D, Zhao X-H, Li Z, et al. Advances in the design and development of SARS-CoV-2 vaccines. Military Medical Research. 2021;8(1):67.

24. Xia S, Duan K, Zhang Y, Zhao D, Zhang H, Xie Z, et al. Effect of an Inactivated Vaccine Against SARS-CoV-2 on Safety and Immunogenicity Outcomes: Interim Analysis of 2 Randomized Clinical Trials. JAMA. 2020;324(10):951–60.

25. Dong Y, Dai T, Wang B, Zhang L, Zeng L-h, Huang J, et al. The way of SARS-CoV-2 vaccine development: success and challenges. Signal Transduction and Targeted Therapy. 2021;6(1):387.

26. Hafiz I, Illian DN, Meila O, Utomo ARH, Susilowati A, Susetya IE, et al. Effectiveness and Efficacy of Vaccine on Mutated SARS-CoV-2 Virus and Post Vaccination Surveillance: A Narrative Review. Vaccines. 2022;10(1):82.

27. Jiang Y, Wu Q, Song P, You C. The Variation of SARS-CoV-2 and Advanced Research on Current Vaccines. Front Med (Lausanne). 2022;8.

28. Sadarangani M, Marchant A, Kollmann TR. Immunological mechanisms of vaccine-induced protection against COVID-19 in humans. Nature Reviews Immunology. 2021;21(8):475–84.

29. Haque A, Pant AB. Efforts at COVID-19 Vaccine Development: Challenges and Successes. Vaccines. 2020;8(4):739.

30. Dong Y, Dai T, Wei Y, Zhang L, Zheng M, Zhou F. A systematic review of SARS-CoV-2 vaccine candidates. Signal Transduction and Targeted Therapy. 2020;5(1):237.

31. Chumakov K, Avidan MS, Benn CS, Bertozzi SM, Blatt L, Chang AY, et al. Old vaccines for new infections: Exploiting innate immunity to control COVID-19 and prevent future pandemics. Proceedings of the National Academy of Sciences. 2021;118(21):e2101718118.

32. Chaudhary JK, Yadav R, Chaudhary PK, Maurya A, Kant N, Rugaie OA, et al. Insights into COVID-19 Vaccine Development Based on Immunogenic Structural Proteins of SARS-CoV-2, Host Immune Responses, and Herd Immunity. Cells. 2021;10(11):2949.

33. Min L, Sun Q. Antibodies and Vaccines Target RBD of SARS-CoV-2. Frontiers in Molecular Biosciences. 2021;8.

34. Al Kaabi N, Zhang Y, Xia S, Yang Y, Al Qahtani MM, Abdulrazzaq N, et al. Effect of 2 Inactivated SARS-CoV-2 Vaccines on Symptomatic COVID-19 Infection in Adults: A Randomized Clinical Trial. JAMA. 2021;326(1):35–45.

35. Kaplan RM, Milstein A. Influence of a COVID-19 vaccine’s effectiveness and safety profile on vaccination acceptance. Proceedings of the National Academy of Sciences. 2021;118(10):e2021726118.

36. Beatty AL, Peyser ND, Butcher XE, Cocohoba JM, Lin F, Olgin JE, et al. Analysis of COVID-19 Vaccine Type and Adverse Effects Following Vaccination. JAMA Network Open. 2021;4(12):e2140364–e.

